# Optimisation of radiographic visibility and wear detection of total knee arthroplasty inlays using radiopaque markers

**DOI:** 10.1101/2025.06.27.25330271

**Authors:** Crystal Kayaro Emonde, Max-Enno Eggers, Klaas Maximilian Heide, Florian Pape, Max Marian, Christof Hurschler, Max Ettinger, Berend Denkena

**Affiliations:** Laboratory for Biomechanics and Biomaterials, Department of Orthopaedic Surgery, Hanover Medical School - Annastift DIAKOVERE, Anna-von-Borries-Strasse 1-7, 30625 Hannover, Germany; Institute of Production Engineering and Machine Tools, Leibniz University Hannover, An der Universität 2, 30823 Garbsen, Hannover, Germany; Institute of Machine Design and Tribology, Leibniz University Hannover, An der Universität 1, 30823 Garbsen, Hannover, Germany; Department of Mechanical and Metallurgical Engineering, School of Engineering, Pontificia Universidad Católica de Chile, Vicuña Mackenna 4860, 6904411 Macul, Región Metropolitana, Chile; Department of Orthopaedic Surgery, University of Oldenburg, Georgstraße 12, 26121 Oldenburg, Germany

**Keywords:** wear marker, micro-milling, UHMWPE, total knee arthroplasty, X-ray, radiopacity

## Abstract

Wear of the inlay in total knee arthroplasty (TKA) contributes to implant failure and the need for revision surgery. *In vivo* wear assessment is challenging owing to the radiolucency of the inlay in standard radiographs. This study aimed to investigate the basic feasibility of integrating radiopaque X-ray markers on standard inlays to enhance their radiographic visibility and enable wear evaluation.

Preliminary experiments identified optimal process parameters for micro-milling cavities on ultra-high molecular weight polyethylene (UHMWPE). A total of 450 parameter combinations were evaluated, with burr formation serving as the quality criterion. A process chain, comprising pre-contouring, micro-milling, filling cavities with radiopaque composite, and final contouring, was developed for inlay production. Eleven inlays with varying marker alignments, orientations, and geometries were manufactured, featuring grooves (≤0.8 mm wide) and holes (diameter = 1.6 mm), both 1 mm deep. Three HDPE + BaSO₄ composites (10, 20, and 30 wt.% BaSO₄) were formulated and assessed for radiopacity per ASTM F640-20. Final marker cavities were filled with HDPE + 20 wt.% BaSO₄ via pellet extrusion. The inlays were positioned in a phantom knee setup and radiographed in the anteroposterior view. Projected markers were evaluated based on edge visibility, measurability, homogeneity, and obscuration by the implant.

None of the parameter combinations resulted in burr-free cavities, indicating an unstable five-axis process. X-ray images revealed that grooves aligned in the X-ray direction and drilled holes exhibited the best visibility for wear markers. Pin-on-plate tribological experiments revealed that BaSO₄ addition to pure HDPE reduced its CoF from 0.25 to 0.1, reaching a value comparable to UHMWPE (0.15), while also enhancing wear resistance.

This study demonstrated the feasibility of integrating wear markers on standard TKA inlays by micro-milling cavities at different positions and orientations on the inlay surface and filling them with a radiopaque composite. Further research is required to optimise process parameters and investigate marker wear.

## 1. Introduction

In 2023, 155,859 primary knee arthroplasties were performed in Germany [1]. Although improved inlay materials have decreased total knee arthroplasty (TKA) revision rates, gross numbers of revision have remained high. That same year (2023), 15,931 revision procedures were also carried out, with revision rates of 4.5% and 5.2% reported at 5 and 9 years, respectively, after implantation [1]. According to the data from major arthroplasty registries worldwide, the main causes of TKA revisions are distributed as follows: aseptic loosening (Germany: 21.6%, Australia: 21.3%; Japan: 34.3%, UK: 31.8%), infection (Germany: 15%, Australia: 27.9%; Japan: 47.9%, UK: 13.5%), instability (Germany: 9.1%, Australia: 10.4%; Japan: 9.4%, UK: 17%), and wear of the inlay component (Germany: 4.9%, Australia: 1.4%; Japan: 6.3%, UK: 13.8%) [1–4].

Wear of the TKA polyethylene tibial inlay component is a cause of late implant failure that compromises the implant’s long-term success and necessitates revision surgery. Studies have reported that younger patients aged ≤ 55 years are at a higher risk of TKA wear due to factors such as higher activity levels and a greater prevalence of obesity [5,6]. Moreover, the wear of the inlay contributes to other mechanisms of TKA failure, including osteolysis and eventual aseptic loosening [7,8]. The German arthroplasty registry reported that in 24% of the TKA revision cases performed in 2023, only the inlay component had been revised [1]. Clinically, TKA patients are routinely monitored radiographically using standard X-ray imaging to assess the implant for malalignment, wear, and early signs of loosening [9]. During these procedures, X-rays of the implanted knee are acquired in standard anteroposterior (AP) and lateral views [10,11]. One of the major limitations of the radiographic evaluation of TKA is the radiolucency of the polymeric inlay component, which renders it invisible in a standard X-ray image [12–14]. Therefore, the assessment of inlay wear is estimated from the change in the relative distance between the visible metallic femur and tibia components, using computer-assisted measurement. More recently, advanced 2D-to-3D matching techniques, such as radiostereometric analysis (RSA), have been investigated [15–17]. However, retrieval analysis remains the primary method to evaluate inlay wear [18,19].

Several studies have investigated concepts to augment the radiographic visibility of the inlay component primarily to evaluate positioning and malalignment *in vivo*. Zaribaf et al. enhanced the radiopacity of the entire inlay component by immersing it in Lipiodol, an iodine-based contrast oil, at an elevated temperature for up to 24 hours, causing the oil to diffuse into the polymer [12]. However, the main challenge with this technique was that the radiopacity reduced over time, compromising its long-term applicability. Other studies incorporated radiopaque metallic markers in the form of tantalum beads inside the inlay to aid migration and dislocation assessment [13,20]. Some commercially available knee arthroplasties, such as Zimmer Biomet’s Oxford Partial Knee, are equipped with metal markers in the form of wires and balls, that have aided in the assessment of bearing dislocation and fracture from standard radiographs [21,22]. However, metal markers pose the risk of inducing fracture of the inlay due to the mismatch in mechanical properties of the polymer and the metal [23].

This study explores the feasibility of incorporating radiopaque markers on the inlay surface to enhance visibility and enable the evaluation of inlay wear. By strategically positioning these markers at the inlay zones most prone to wear, it is hypothesised that inlay wear can be better predicted and visualised compared to pure reference materials. Microstructures in the form of grooves with a maximum width of 0.8 mm and holes with a diameter of 1.6 mm, both with a depth of 1 mm, are machined on the inlay surface and later filled with a radiopaque composite via an extrusion process. Eleven marker geometries are investigated focusing on the specific process parameters necessary to micro-mill these structures on ultra-high molecular weight polyethylene (UHMWPE).

In contrast to conventional milling in the macro-scale, micro-milling is dominated by simultaneous shear and ploughing forces. Therefore, this process needs further analysis, particularly for the machining of polymers. Moreover, the literature on micro-milling of UHMWPE is very limited, which makes it difficult to select suitable process control variables. However, preliminary investigations into the micro-machining of polymers have shown promising results [24]. Kuram et al. reported successful machining of polypropylene (PP) [25]. Adeniji et al. also demonstrated that burr formation plays a crucial role in the component quality when micro-milling polymers [26].

The wear marker material employed in this study comprises a mixture of high-density polyethylene (HDPE) and BaSO_4_, a contrast agent widely used in clinical radiography to visualise the gastrointestinal tract. It is also present in some bone cements and biodegradable implants for the same purpose [14]. Three composites with varying concentrations of BaSO_4_ and HDPE are formulated and their radiopacity is evaluated based on ASTM F640-20, a standard used for radiopacity testing for medical use.

To evaluate the quality of the final wear markers, a weighted scoring system is developed based on four criteria: edge visibility, marker measurability, homogeneity of the fill, and marker obscuration by the implant. This approach aids the selection of the most optimal wear marker geometries that can accurately and reliably estimate wear. Additionally, the tribological properties of the marker composites are evaluated to investigate any differences with respect to the bulk material. The fact that wear markers exhibit uniform wear consistent with the bulk inlay material is established as an evaluation criterion to provide an accurate representation of actual wear occurring between the femur component and the inlay.

## 2. Materials and Methods

### 2.1 Preparation of radiopaque wear marker material

For the radiopaque wear marker material, three different composites were prepared by blending BaSO₄ powder (Sigma Aldrich Chemie GmbH, Germany) at mass compositions of 10, 20, and 30 wt.% with HDPE pellets (HDPE 76303, SK Global Chemical, China)^1^. Initially, PE1000 (UHMWPE) was considered as the base material to match the bulk inlay material. However, due to challenges processing UHMWPE [27], HDPE 76303, with a density of ρ = 0.956 g/cm³ and a Vicat softening point of T = 123 °C, was selected as the closest alternative. BaSO_4_ was used as a radiopacifier. HDPE and BaSO_4_ were mixed using a laboratory twin-screw extruder operating at a temperature range of 190-200 °C, and a screw speed at 200–300 rpm. Three composites were thus produced: HDPE with 10, 20, and 30 wt.% BaSO_4_, respectively, all of which comprised cylindrical pellets with a maximum size of < 3 mm.

### 2.2 Inlay and wear marker design

Standard inlays (75 mm × 50 mm × 20 mm) were designed and manufactured from non-crosslinked PE1000 by milling. The inlays represented a typical geometry and were adapted from the commercially available Stryker Triathlon cruciate-retaining (CR) size 6 inlay. All milled inlays had the same thickness. Eleven marker patterns primarily comprising grooves and drilled holes were then conceptualised to find suitable geometries for the intended purpose. This procedure was based on the work of Denkena et al. [24]. The general cavity depth for the microstructures was based on reported clinical linear wear rates of standard inlays. On average, linear wear of a TKA inlay is estimated at 0.5 mm in the first year, after which it stabilises to 0.1–0.2 mm in the following years [28]. Thus, the specified depth of d_c_ = 1 mm was found sufficient for the present study. The milled cavities had a width of w_c_ = 0.8 mm and a length of l_c_ = 4 mm, while the holes had a diameter of D_h_ = 1.6 mm.

The structures were integrated into the free-form surface of the implant surface. Two cavity configurations were selected, whereby the cavities were either aligned parallel to the surface or parallel to the backside of the inlay (Figure 1). In the first case (Figure 1a), the cavity depth d_c_ remained constant over the entire length, whereby the actual length l corresponded to the planned target length l_c_. In contrast, the depth d_c_ of the cavity aligned parallel to the backside of the inlay varied along the groove (Figure 1b). In this case, the mean depth was adapted to the target value. A special case occurred when the angle α between the surface and the bottom of the cavity was excessive (Figure 1c), In this case, it would not be possible to fully achieve the target length l_c_.

**Figure 1:**
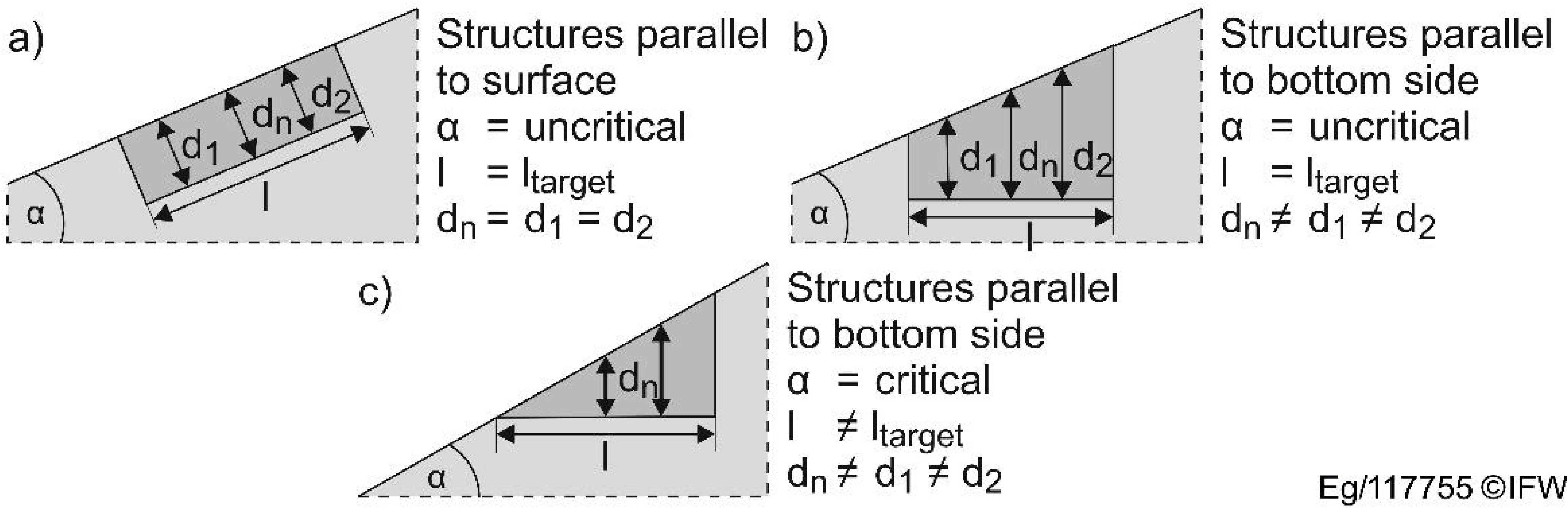
Structure alignment a) parallel to surface, b) parallel to backside c) with critical angle.

Eleven different marker geometries and patterns (P) were investigated (Figure 2). The structures comprised grooves and/or drilled holes at different positions and orientations across the articulating inlay surface. The structures were aligned along a horizontal reference line that extended over the medial and lateral condyles and marked the zones of the inlay most susceptible to wear [29,30].

**Figure 2:**
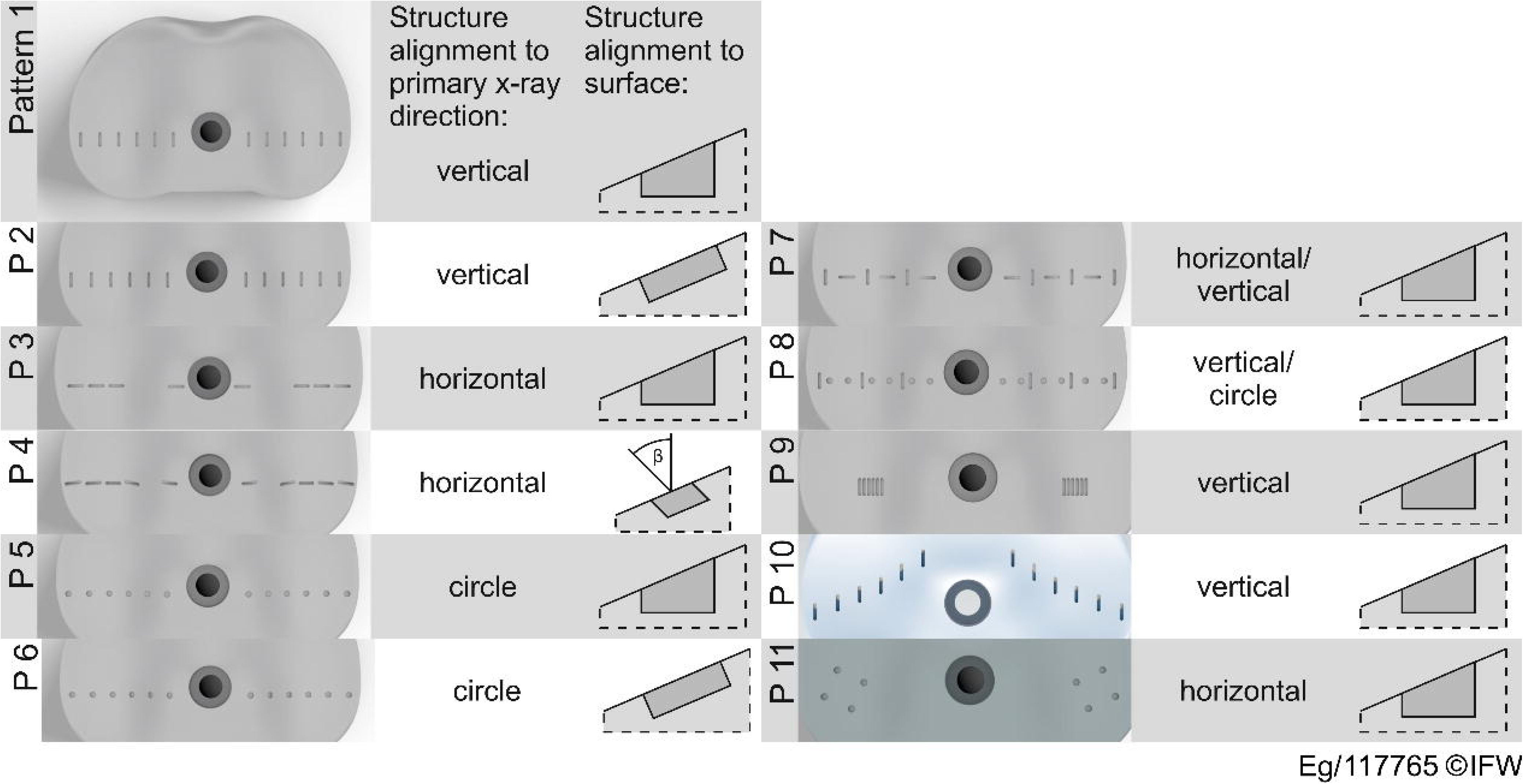
Proposed patterns for the 11 wear marker patterns.

The reference structure (pattern 1) comprised vertical grooves aligned parallel to the backside of the inlay, spaced 5 mm apart. This spacing would ensure clear radiographic visibility of the individual markers as it significantly exceeded the resolution of conventional X-ray devices (0.05–0.17 mm). All other patterns had a marker spacing in the range of 1– 5 mm, except pattern 9, whose inter-marker distance was 0.2 mm, which was marginally above the upper resolution limit. Furthermore, a distinction was made between patterns that were parallel to the backside of the inlay-patterns that were aligned perpendicular to the inlay surface (normal direction) and pattern 4, in which the structures were inclined at β = 30° in order to assess how the angulation of the markers influences the X-ray projection. In addition to different orientations of the grooves (horizontal and vertical to the X-ray direction), different structures (grooves and holes) were investigated in different combinations. The structures in samples 10 and 11 were offset anteriorly to investigate the effect of the marker positioning relative to the X-ray source. Together, these proposed patterns were designed to provide a comprehensive analysis of the visibility and functionality of potential X-ray markers and could be used as a basis for further investigations.

### 2.3 Inlay manufacturing

#### Initial parameter studies for micro-milling

Initial process parameters were selected based on published literature and previous studies [24] (Table 1). To determine suitable process parameters to produce the structures, preliminary experiments were conducted using milling tools 6 and 7 on a flat surface (Table 2). The trials were performed according to a full factorial experimental design with three repetitions per case.

**Table 1:** Process control variables used for the parameter study.

| Tool diameter | Cutting depth | Tooth feed | Cutting speed |
| --- | --- | --- | --- |
| [mm] | [mm] | [mm] | [m/min] |
| 0.5 | 0.075 | 0.0050 | 10 |
| 0.6 | 0.100 | 0.0125 | 15 |
|  | 0.125 | 0.0200 | 20 |
|  | 0.150 | 0.0600 | 25 |
|  | 0.200 | 0.1000 | 30 |

**Table 2:** Specifications of milling tools for contouring and micro-structuring.

| Nr. | Tool diameter<br>[mm] | Cutting edges<br>[-] | Tool type<br>[-] |
| --- | --- | --- | --- |
| 1 | 10.0 | 1 | End mill |
| 2 | 10.0 | 2 | Ball mill |
| 3 | 6.0 | 1 | End mill |
| 4 | 2.0 | 1 | End mill |
| 5 | 1.0 | 1 | End mill |
| 6 | 0.6 | 3 | End mill |
| 7 | 0.5 | 3 | End mill |

The quality of the micro-milled cavities was evaluated based on the degree of burr formation, as this was critical for the filling with the radiopaque composite. For this purpose, images were obtained using a reflected light microscope (Keyence VHX-5000).

#### Evaluation of micro-milled cavities

Burr formation served as the primary criterion for evaluating the process control variables after micro-milling the marker cavities. The evaluation method has been previously described in [24]. Based on the external appearance of the samples, burr formation was classified into five categories:

1. **Burr-free:** No burr or only isolated threads
2. **Few threads:** Fine threads in small quantities
3. **Wavy:** Wavy tabs on the groove edges with isolated threads
4. **Multiple threads:** Coarse threads in large quantities on the groove edge, some of which are protruding inwards or outwards
5. **Clogged groove:** Heavy burr formation with additional partial filling of the groove

This classification based on the microscopy images allowed for a systematic evaluation of the factors influencing burr formation and served as a basis for optimising the process parameters.

#### Process chain for inlay manufacturing

The process chain for manufacturing the marker-integrated TKA inlays is summarised in Figure 3. First, conventional milling of the contour was carried out with an allowance of 0.3 mm for finishing. Next, the microstructures for the wear markers were precisely machined by micro-milling. The cavities were subsequently filled with the marker material through an extrusion process. Lastly, subtractive end contouring was performed to create the final geometry of the inlay.

**Figure 3:**
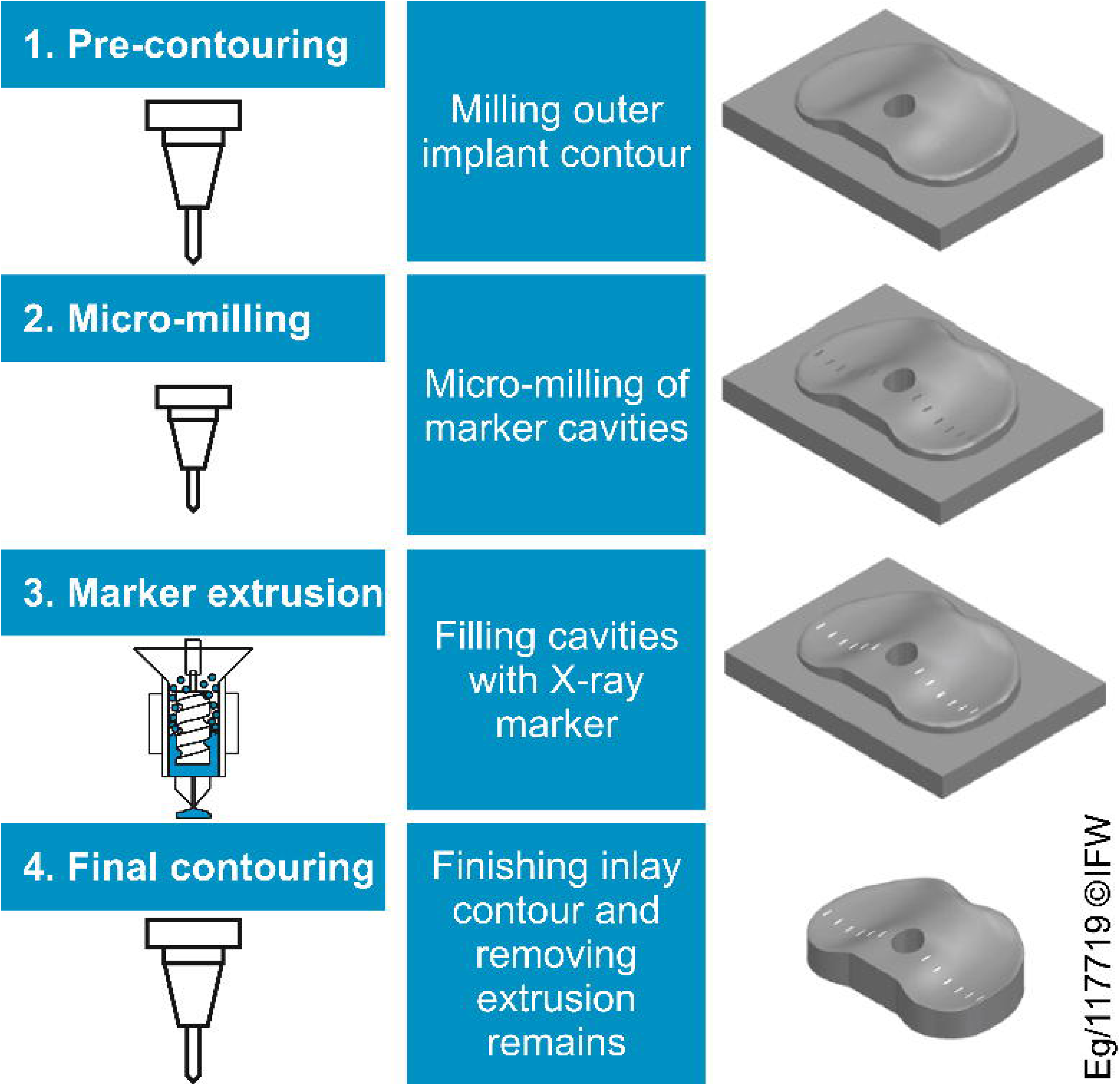
Process chain for the production of marker-integrated inlays.

UHMWPE in the form of PE1000 (ADS Drehservice GmbH) was used as the starting material for the inlays. The polymer was a block measuring 100 mm × 75 mm × 30 mm with a density of *ρ* = 0.93 g/cm³ and a Vicat softening point of T = 130 °C.

The subtractive processing steps were performed on a five-axis milling centre (Ultrasonic 10, DMG Mori AG). The machine had a maximum spindle speed of 40,000 rpm and a maximum torque of 2 Nm. The milling tool specifications are listed in Table 2.

The process parameters determined in the preliminary tests were adopted for the production of micro-cavities on the standard inlays. After micro-milling on the standard inlay surface, burr formation was reanalysed. For systematic evaluation, the microstructures were divided into the following categories:

i. Grooves aligned parallel to the backside of the inlay
ii. Grooves aligned parallel to the inlay surface (normal vector)
iii. Grooves inclined at an angle 30°
iv. Holes aligned parallel to the backside of the inlay
v. Holes aligned parallel to the inlay surface (normal vector)

The generated micro-cavities were filled with the radiopaque marker material^2^. Melting and extrusion were carried out with a pellet extruder (V4, Mahor.xyz, Spain) using a 0.6 mm nozzle at a processing temperature of T = 180 °C. Owing to its free-form surface, the inlay was manually positioned under the extruder during filling.

### 2.4 Quantitative determination of wear marker radiopacity

Ten flat samples (55 mm × 15 mm × 3 mm) were produced from pure UHMWPE, pure HDPE, and the three radiopaque composites containing 10, 20, and 30 wt.% BaSO₄. Of the ten samples, five were used for radiopacity evaluation, while the other five were used to test the tribological properties. The samples were compression moulded at a process temperature range of 190–200 °C using a hand lever press and milled to the final contour to achieve a surface finish similar to that of the inlay. The surface roughness was determined in accordance with ISO 4287 (Table 3.

**Table 3:** Surface roughness R_a_ and R_q_ of the samples used for the determination of radiopacity and tribological.

| Sample material | Arithmetic Average<br>Roughness $R_a$[$\mu\text{m}$] | Root Mean Square<br>Roughness $R_q$[$\mu\text{m}$] |
| --- | --- | --- |
| UHMWPE | 1.682 | 2.043 |
| HDPE + 10 wt.% BaSO<sub>4</sub> | 0.981 | 1.227 |
| HDPE + 20 wt.% BaSO<sub>4</sub> | 1.017 | 1.251 |
| HDPE + 30 wt.% BaSO<sub>4</sub> | 0.853 | 1.063 |

The radiopacity values of the samples were obtained by converting their greyscale values within a 0–255 range into an equivalent thickness of aluminium in accordance with ASTM F640-20. For this purpose, a reference aluminium 1100 step wedge, with 21 steps with 1 mm graduations per step, was positioned beside the samples during X-ray image acquisition. Additionally, a sample SI containing micro-milled holes filled with the 20 wt.% BaSO₄ wear marker composite was positioned alongside the samples to assess the visibility of the wear markers within a standard inlay (Figure 4).

**Figure 4.**
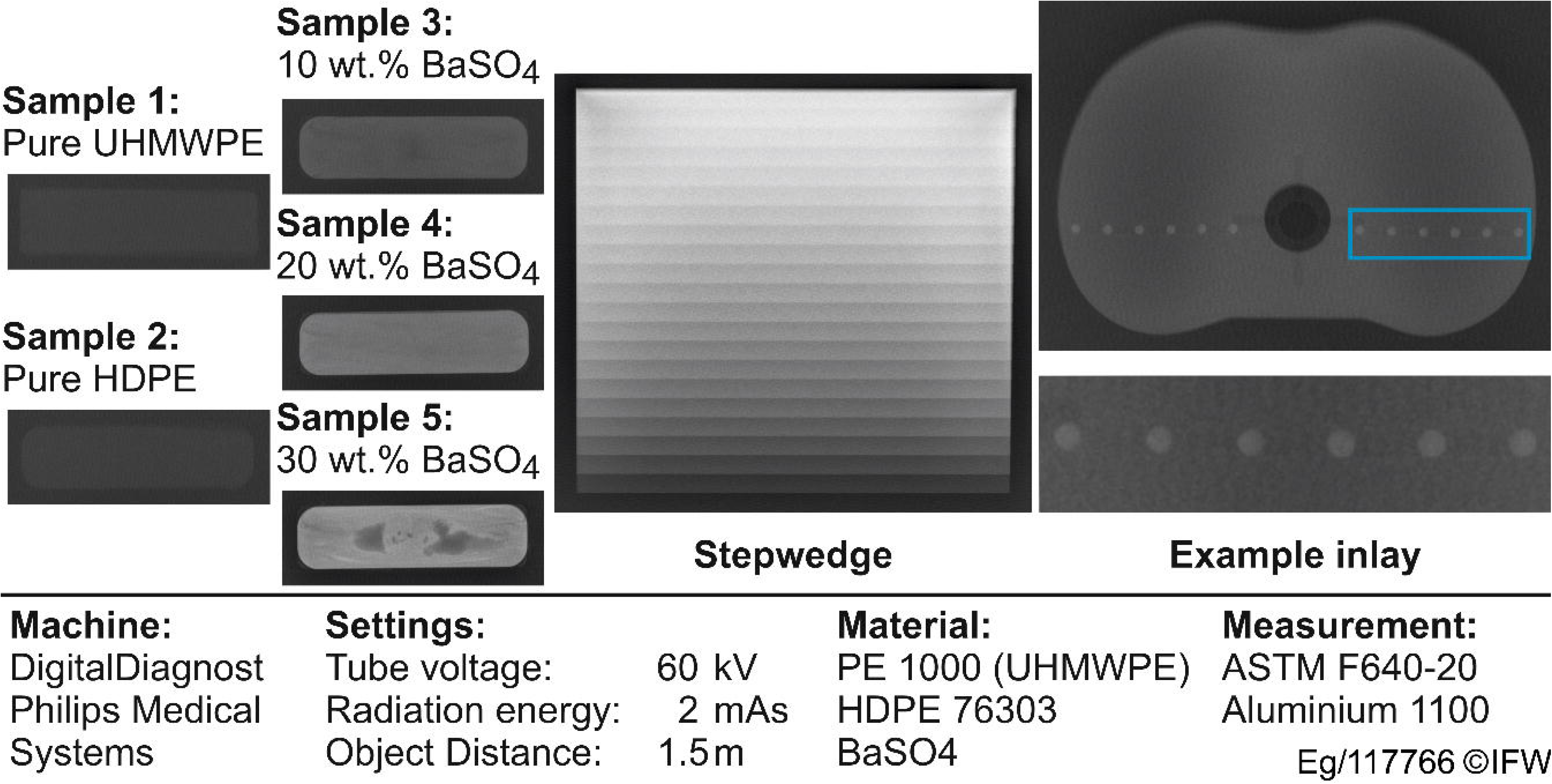
X-ray radiographs of the samples, stepwedge and a marker-integrated inlay sample.

Digital X-ray radiographs of the samples were obtained using a Philips Medical Systems X-ray machine (DigitalDiagnost, Philips Medical Systems, Netherlands). The samples were imaged in top view at 60 kV and 2 mAs and a source-to-detector distance of 1.5 m.

For each step of the aluminium step wedge, a uniformly sized rectangle was placed within the step and the greyscale value was determined using image analysis software (ImageJ, version 1.54, USA) on the unaltered X-ray image. The recorded greyscale values and their equivalent steps were then fitted by linear regression as described by Nomoto et al. [31]. For each sample, greyscale measurements were conducted three times, at different positions and subsequently averaged. This was also done for the image background. The radiopacity of the background was then subtracted from that of the samples to determine the actual radiopacity of the samples.

### 2.5 Tribological assessment

For tribological experiments, a pin-on-plate tribometer (Tribotechnic, France) was used. In this setup, the five samples were successively mounted on an oscillating plate and tested under sliding motion against a fixed Al₂O₃ ball (D_Al2O3_ = 6 mm) under a defined load of 2 N. The setup has been detailed in a previous study by Pape et al. [32]. The test parameters included a stroke length of 2 mm, a sliding velocity of 8 mm/s, and a total sliding distance of 300 m. The experiments were conducted under dry and ambient conditions, with frictional force being continuously recorded throughout the test. After test completion, the surface was analysed using a laser scanning microscope (VK-X200, Keyence, Japan). The profiles of the samples were also plotted along the wear track to study the tribological characteristics of the samples.

### 2.6 *In vitro* analysis of radiographic wear marker visibility

Each of the filled marker-integrated inlays was successively introduced between TKA components (size 6, Stryker Triathlon CR, Stryker European Operations B.V., Netherlands) fitted on the phantom knee setup and then imaged (Figure 5). The procedures for positioning and X-ray acquisition used have been previously described [16]. For each inlay, radiographs were acquired in the AP view at typical knee imaging flexion angles 0° and 30°, using a ceiling-mounted digital X-ray system, with imaging parameters of 1.4 mAs and 60 kV, and a source-to-detector distance (SDD) of 1.5 m. Additionally a calibration sphere (*D*_s_ = 25 mm) was positioned within the radiographic view. The final radiographs were uniformly enhanced in the X-ray software before saving.

**Figure 5.**
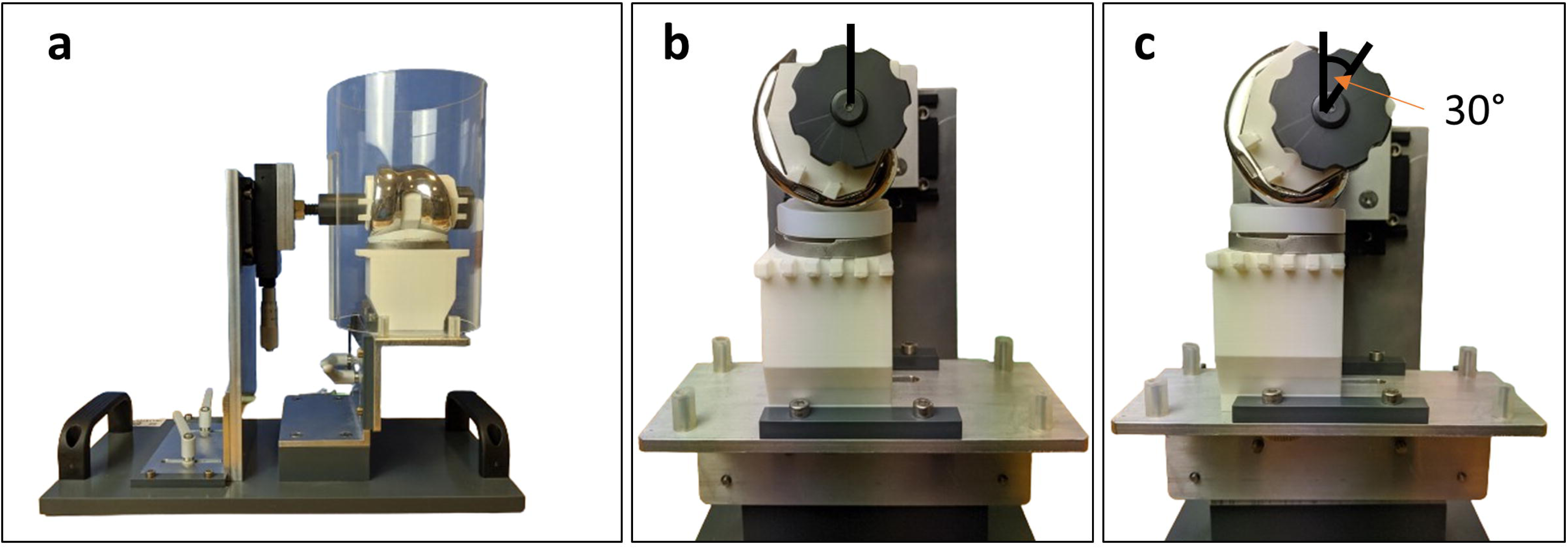
(a) Front view. Lateral views illustrating the femur component at (b) 0° and (c) 30°.

### 2.8 Wear Marker Evaluation

The next step involved evaluating the radiographic quality of the filled markers based on the acquired radiographs. A weighted scoring system based on four criteria was created to rank the filled markers and to identify the markers that would be most optimal for use as potential inlay wear markers. These criteria included edge definition, measurability, homogeneity, and marker obscuration by the implant. For each inlay, the evaluation was performed on all visible markers, and the average value was used for final scoring per category. All radiographs acquired for each inlay were evaluated for each criterion. The weight of each criterion was assigned based on its importance to the determination of inlay wear (Table 4).

**Table 4.** Summary of scoring criteria for wear marker quality evaluation.

| Criteria | Scale | Weight | Max. points |
| --- | --- | --- | --- |
| Edge definition | 1–4 | x3 | 12 |
| Measurability | 1–3 | x2 | 6 |
| Homogeneity | 1–3 | x2 | 6 |
| Marker obscuration | 1–3 | x1 | 3 |
| Weighted score |  |  | **27** |

#### Edge definition

This criterion comprised a four-point scale. The highest score of 4 represented an ideal case scenario, where markers had well-defined edges (Figure 6). Edge definition was assigned the highest weight as the evaluation of other criteria, such as homogeneity and measurability, was dependent on the clarity of the edges.

**Figure 6.**
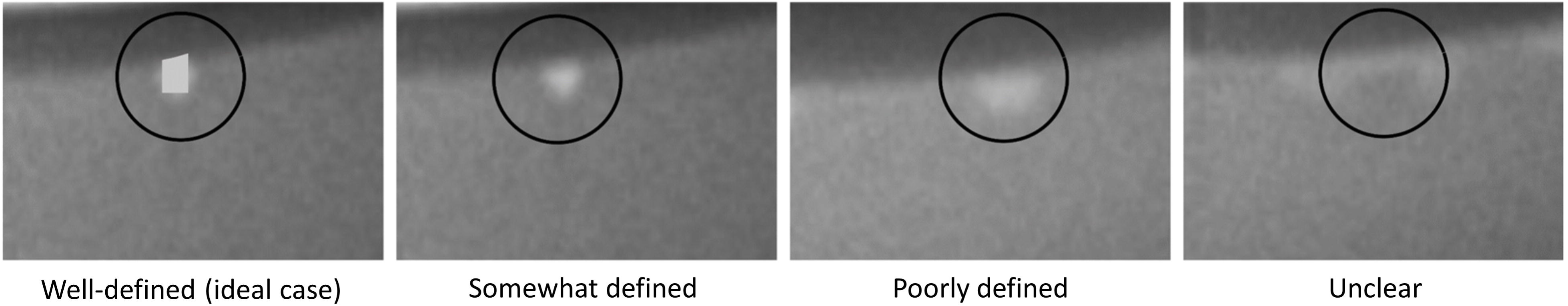
Scale description of the four-point edge definition evaluation criterion.

#### Measurability

Measurability was defined as the ability to quantify the marker dimensions (depth and width) from the radiograph. The calibration sphere was used as a reference for scale. Automatic measurement of the marker dimensions was not directly possible as there was no clear distinction of the marker edges from the bulk inlay. Thus, greyscale variation was used for this measurement. However, this required the marker edges to be moderately visible (edge definition score of 3). For marker width determination, greyscale measurements were taken along the widest section of the marker and the peripheral inlay region. Based on a greyscale distribution of 0–100%, where 0% represents the minimum possible greyscale value of 0 and 100% represents the maximum value of 255, the pixel-to-pixel intensity within the marker region was observed to have a maximum variation of 7.84%. Thus, a pixel-to-pixel greyscale variation exceeding this threshold was considered a transition from the marker to the bulk inlay or to the femur component (Figure 7).

**Figure 7.**
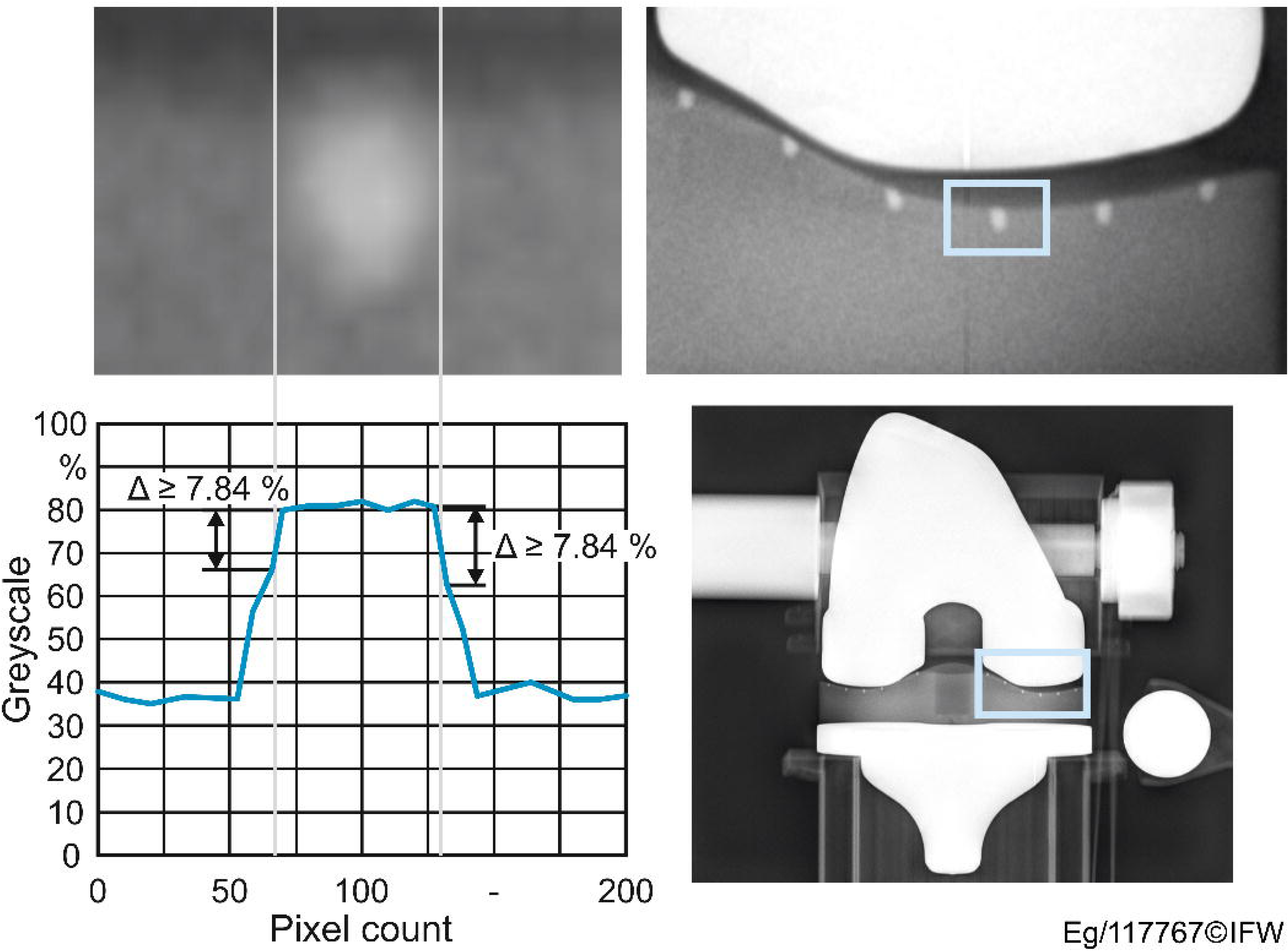
Greyscale analysis on a marker indicating the marker region and transition from marker to inlay.

Similarly, for marker depth measurement, greyscale measurements were taken from the top to the bottom edges of the marker. To ensure that the marker depth corresponded to the 1 mm depth defined in the process chain, measurements were taken from the centre of the bottom edge, proceeding upwards. For accurate marker depth measurements, it was essential that the top edge of the marker is distinguished from the radiopaque femur component even at contact. The measured values were compared with the true geometries from the planned 3D-model and the deviations (mean error) were used to rank the marker measurability.

#### Homogeneity

Homogeneity was assessed to evaluate the quality of the fill using greyscale values as the measurement criterion. This evaluation was not possible for markers with unclear edges (edge definition score of 1). For groove markers, a square measuring 5 pixels × 5 pixels (0.1 mm/pixel) was positioned at the centre of the marker to measure the greyscale value. The grooves had a cross-sectional area of approximately 0.8 mm × 1 mm. For the hole markers (D_h_ = 1.6 mm), a square measuring 6 pixels × 6 pixels (0.1 mm/pixel) was used for measurement. The analysed areas were therefore smaller than the maximum cross-sectional area of both the hole and groove. This minimised errors due to regions outside the marker being included in the measurement. The result was an averaged greyscale value with a specified standard deviation for all markers evaluated per inlay. The standard deviation of the greyscale was used to assess the homogeneity of the greyscale distribution.

#### Marker obscuration

Marker obscuration was assessed to determine how clearly the markers could be visualised separately from the femur at both flexion angles. A three-point scale was used to quantify the extent of marker obscuration, with the highest score of 3 indicating that all markers were clearly distinguishable without any obscuration from the articulating femur.

A summary of the score descriptions for the evaluation criteria is provided in (Table 5).

**Table 5.**
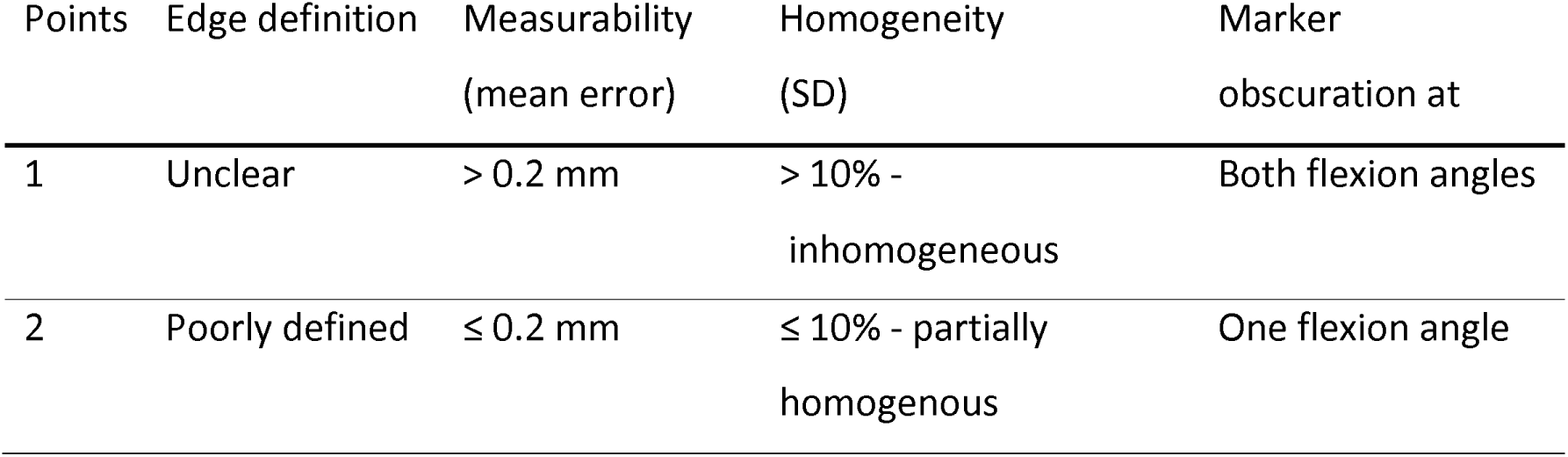
Scale descriptions of the four criteria for wear marker quality evaluation.

## 3. Results

### 3.1 Process parameters for micro-milling and inlay manufacturing

#### Initial parameter studies

Burr-free micro-cavities were not achieved for any of the process parameter combinations. The average burr class for samples produced by a tool diameter of D_T,1_ = 0.5 mm and D_T,2_ = 0.6 mm was 3.79 and 4.50, respectively, indicating increased burr formation with a larger tool diameter. The lowest burr formation was achieved with a parameter combination comprising a tool diameter of D_T_ = 0.5 mm, cutting depth of a_p_ = 0.075 mm, feed per tooth of f_z_ = 0.03 mm, and cutting speed of v_c_ = 10 m/min, which corresponded to an average burr class of 2.67, characterised by few threads to wavy tabs on the microstructures.

As no suitable regression model for determining optimal process parameters could be derived from the test results, the parameters that resulted in the lowest burr formation were defined as variables for production on the standard inlays (Figure 8).

**Figure 8:**
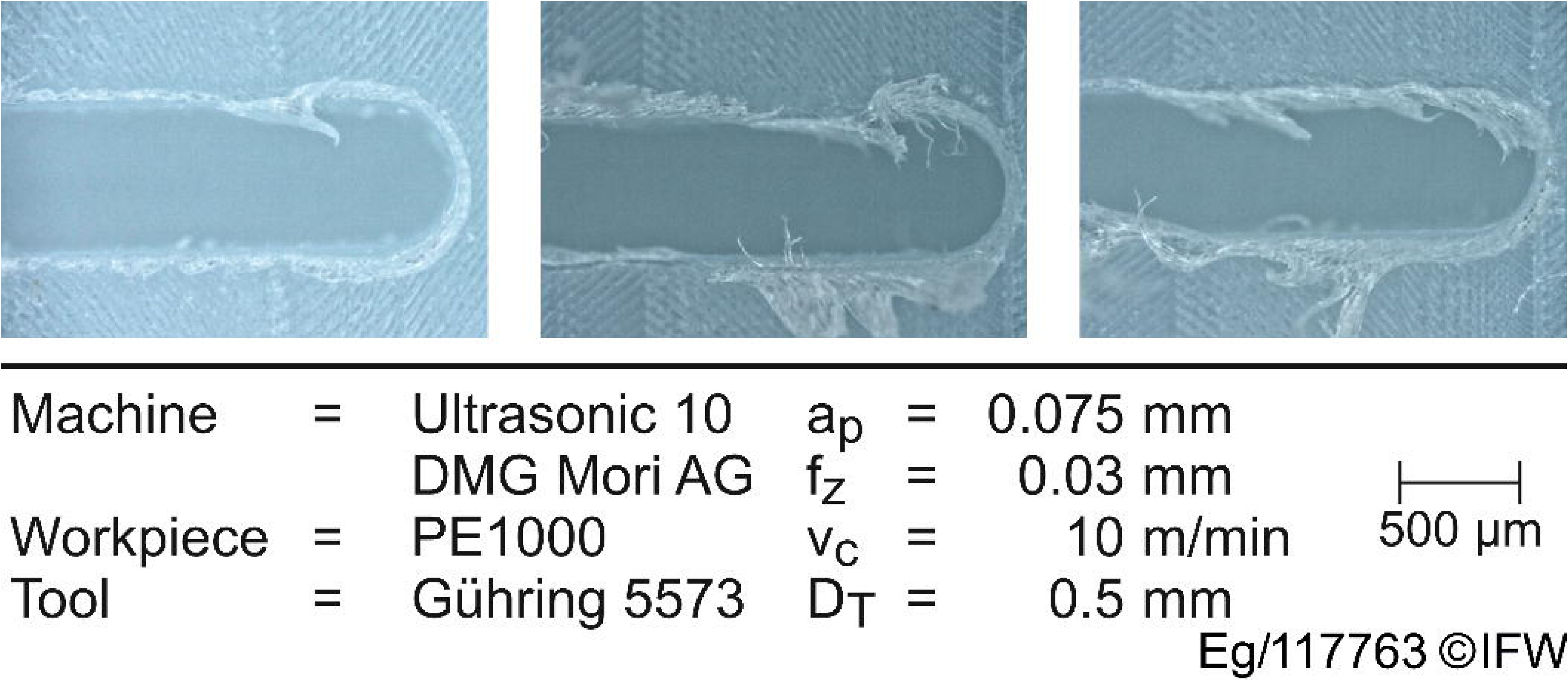
Microscope images of the samples with the lowest average burr formation.

#### Inlay manufacturing

The mean burr classes were determined for each of the five micro-cavity categories (Table 6). There was a noticeable difference in burr formation compared to the parameter study. Burr formation was substantially less in holes than in grooves. The difference between the burr formation for grooves aligned parallel to the backside and those along the normal vector of the surface was marginal in both cases. The grooves inclined at 30° exhibited substantially higher burr formation. Manual removal of burrs was necessary in all cases prior to filling the grooves with the radiopaque marker composite.

**Table 6.**
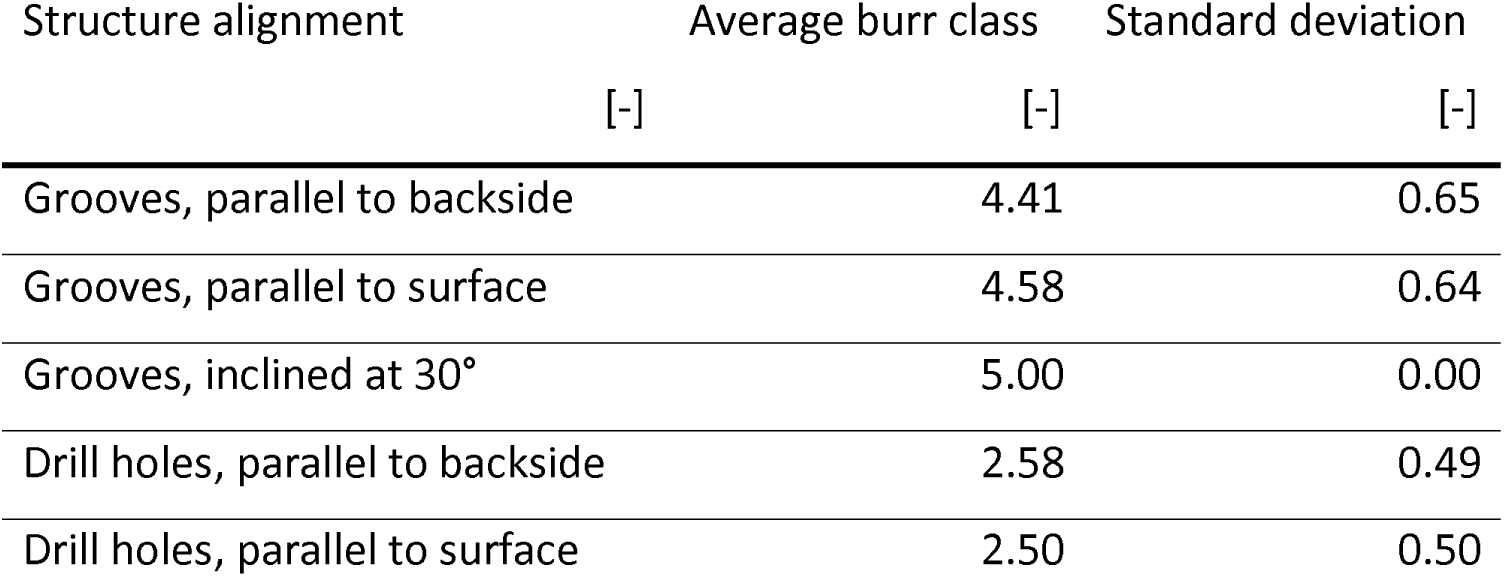
Average burr classes for the milled microstructures on the standard inlays.

| Structure alignment | Average burr class | Standard deviation |
| --- | --- | --- |
| [-] | [-] | [-] |
| Grooves, parallel to backside | 4.41 | 0.65 |
| Grooves, parallel to surface | 4.58 | 0.64 |
| Grooves, inclined at 30° | 5.00 | 0.00 |
| Drill holes, parallel to backside | 2.58 | 0.49 |
| Drill holes, parallel to surface | 2.50 | 0.50 |

During the finishing of the inlay contour, it was observed that some of the filled markers partially dislodged from the cavities. However, no damage was observed to the surrounding material. This phenomenon occurred randomly at different points; thus, no clear conclusions could be drawn about possible irregularities in the manufacturing process.

### 3.2 Evaluation of filled wear markers

In this section, the weighted scores of the filled marker patterns are presented based on the four criteria described previously.

#### Edge definition

Of all the microstructures, the vertical groove markers exhibited the highest edge definition, followed by the holes (P5, P6, P11, and partly P8), which had slightly less defined edges (Figure 9). The lowest edge definition was observed for the horizontal grooves (P3, P4, and partly P7).

**Figure 9.**
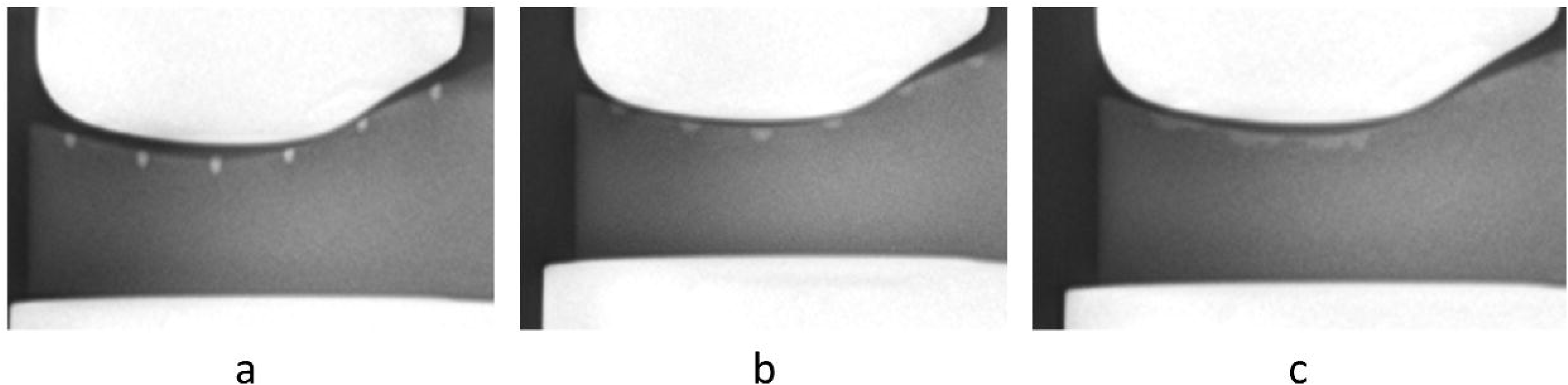
Edge definitions of inlay containing (a) vertical grooves, (b) holes, and (c) horizontal groove markers.

The variation in the edge definition can be described by the Beer’s law of X-ray attenuation

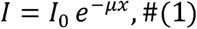

which states that the transmitted X-rays (I_o_) are directly proportional to the material thickness (x) and the attenuation coefficient (*µ*) of the material. In the present case, the attenuation coefficient is constant as the marker composite used in all micro-cavities was the same. However, the incident X-rays traversed varying path lengths (material thicknesses) for the different microstructures. The vertical groove marker, having the longest path length through which the X-rays travels (x = 4 mm), exhibited the highest attenuation. Thus, the markers had the highest visibility and definition, followed by the holes with a thickness x = 1.6 mm and finally the horizontal markers with a thickness x = 0.8 mm (Table 7).

**Table 7.** Average scores for the 11 marker patterns containing groove and holes in varying combinations.

| Pattern | 1 | 2 | 3 | 4 | 5 | 6 | 7 | 8 | 9 | 10 | 11 |
| --- | --- | --- | --- | --- | --- | --- | --- | --- | --- | --- | --- |
| Edge definition | 3.0 | 3.0 | 1.0 | 1.0 | 2.0 | 2.0 | 1.6 | 2.2 | 3.0 | 3.0 | 2.0 |
| Measurability | 2.7 | 2.5 | 1.0 | 1.0 | 1.0 | 1.0 | 1.5 | 1.3 | 2.6 | 2.7 | 1.0 |
| Homogeneity | 2.6 | 2.8 | 1.0 | 1.0 | 3.0 | 3.0 | 1.5 | 2.9 | 2.2 | 2.4 | 3.0 |
| Obscuration | 2.9 | 3.0 | 2.5 | 2.6 | 3.0 | 2.8 | 2.2 | 3.0 | 3.0 | 3.0 | 3.0 |

Additionally, the specified inter-marker distance had no effect on marker visibility. A comparison between markers with the largest and smallest inter-marker spacing (5 mm and 0.2 mm) demonstrated sufficient visibility in both cases (Figure 10). Nevertheless, a marker spacing below the resolution of the X-ray device may result in reduced visibility of the individual markers.

**Figure 10.**
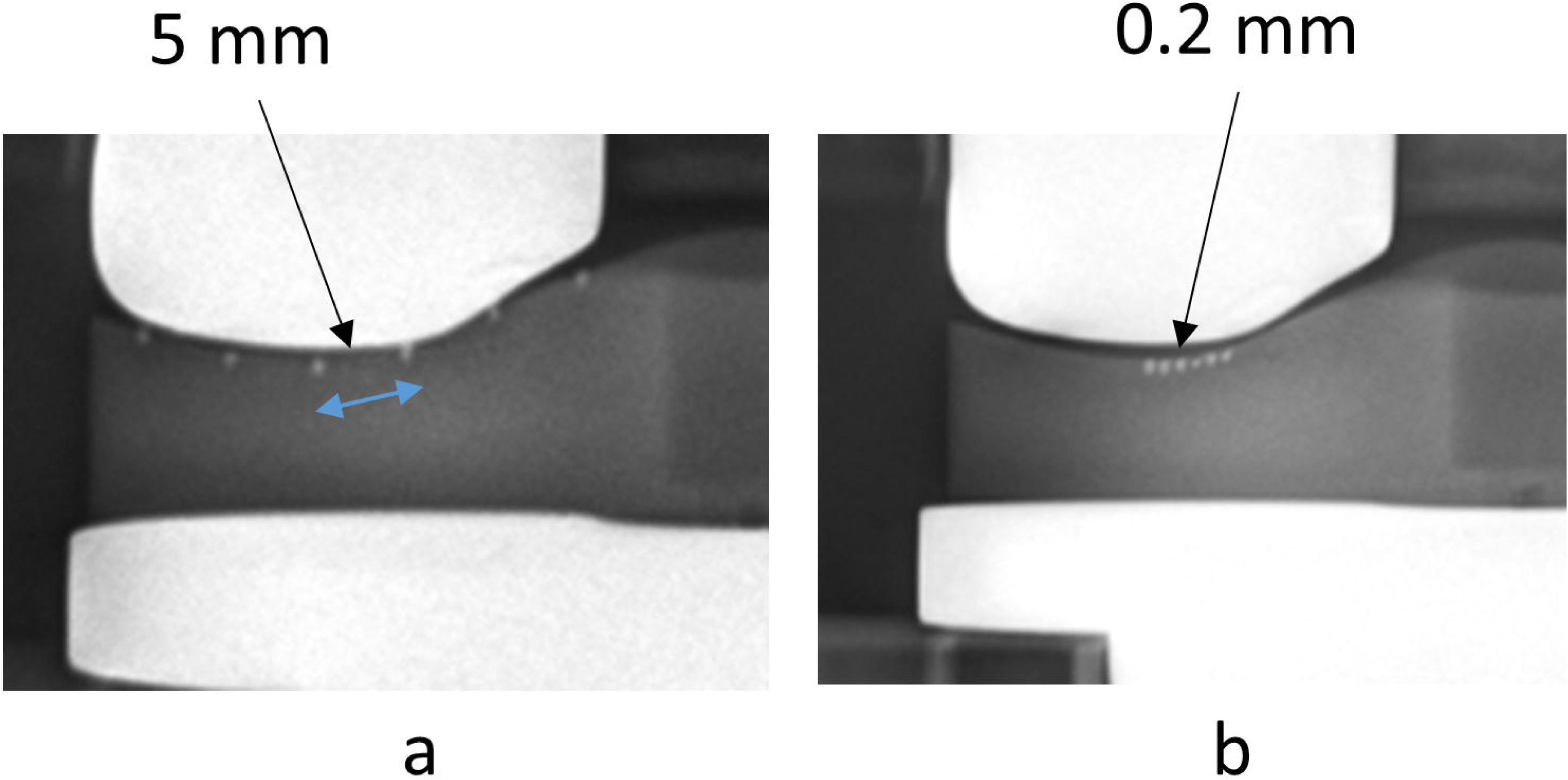
Comparison of wear markers with a) 5 mm and b) 0.2 mm inter-marker spacing.

#### Measurability

Measurability accuracy was highest for the vertical groove markers aligned in the direction of the X-rays (P1, P2, P9, P10), all of which achieved a score of ≥2.5 (Table 7). These microstructures also exhibited the highest edge definition, highlighting the importance of clear edges for precise marker dimension measurement. Another factor that could result in variations in the marker dimensions from those specified in the process chain could be insufficient filling of the microstructures due to insufficient burr removal.

#### Homogeneity

Homogeneity measurements were only possible for markers that had an edge definition score >1.0 (Table 7). The highest homogeneity values were found for the holes (P5, P6, P11, and partly P8) followed by the vertical grooves (P1, P2, P9, P10, and partly P8). The horizontal groove markers (P3, P4, and partly P7), owing to their low edge definition, exhibited the poorest homogeneity scores.

#### Marker obscuration

In the in vitro radiographs on the phantom knee model, the inlay component appeared partially visible. Clinically, this is not the case, and the inlay is entirely radiolucent. From the radiographs, it was observed that most markers remained entirely visible and distinguishable from the high contrast femur component. Nevertheless, there were cases where the markers were partially obscured by the contacting femur component due to positioning.

For each criterion, the final scores were an average of all the markers evaluated on each individual inlay (Table 7***Error! Reference source not found***.).

Once all the criteria were evaluated, the final marker scores were weighted (Table 8). The highest scores (>20) were observed for the vertical grooves, followed by the holes. The horizontal grooves exhibited the lowest scores.

**Table 8.** Cumulative weighted scores for the 11 marker patterns based on the evaluation criteria.

| Pattern | weighted<br>edges | weighted<br>measurability | weighted<br>homogeneity | weighted<br>obscuration | Total<br>score |
| --- | --- | --- | --- | --- | --- |
| 1 | 9.0 | 5.4 | 5.2 | 2.9 | 22.5 |
| 2 | 9.0 | 5.0 | 5.6 | 3.0 | 22.6 |
| 3 | 3.0 | 2.0 | 2.0 | 2.5 | 9.5 |
| 4 | 3.0 | 2.0 | 2.0 | 2.6 | 9.6 |
| 5 | 6.0 | 2.0 | 6.0 | 3.0 | 17.0 |
| 6 | 6.0 | 2.0 | 6.0 | 2.8 | 16.8 |
| 7 | 4.8 | 3.0 | 2.0 | 2.2 | 12.0 |
| 8 | 6.6 | 2.6 | 5.8 | 3.0 | 18.0 |
| 9 | 9.0 | 5.2 | 4.4 | 3.0 | 21.6 |
| 10 | 9.0 | 5.4 | 4.8 | 3.0 | 22.2 |
| 11 | 6.0 | 2.0 | 6.0 | 3.0 | 17.0 |

### 3.3 Evaluation of wear marker composite radiopacity

As anticipated, BaSO_4_ concentration was directly correlated with the radiopacity of the composites (0.03).

Figure 11). The equivalent aluminium thickness (mmAl) of the samples was calculated based on the regression equation (Equation 2) of the aluminium thickness and the corresponding greyscale value of the step wedge:

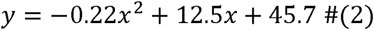

**Figure 11.**
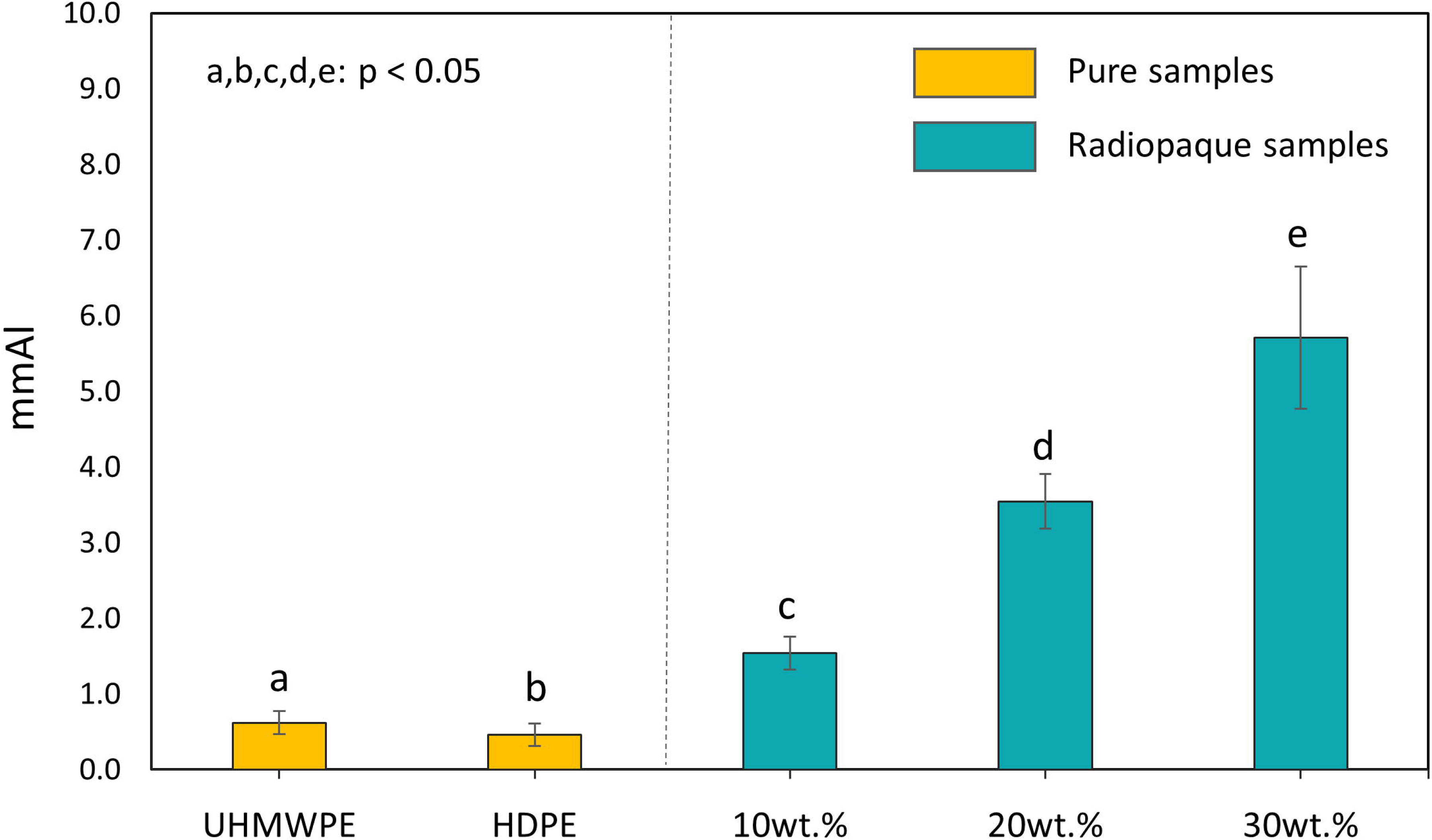
Radiopacity of pure UHMWPE, HDPE, and HDPE+ (10, 20, and 30) wt.% BaSO_4_ composite samples.

Statistically significant differences were observed among all HDPE + (10, 20, and 30) wt.% BaSO₄ composites, with radiopacities of 1.54, 3.54, and 5.71 mmAl, respectively (Welch’s t-test, p < 0.05). Additionally, significant differences were also found between the radiopacities of the pure UHMWPE and HDPE samples (0.62 mmAl vs. 0.46 mmAl, p = 0.03).

### 3.4 Tribological results

The evolution of the coefficient of friction (CoF) over test length is displayed in Figure 12. Of all the five tested samples, pure HDPE exhibited the highest friction, with an initial CoF of 0.18 that increased within the first meters to approximately 0.25, before stabilising. The UHMWPE sample had the second-highest CoF, ranging from approximately 0.14 to 0.15. By incorporating BaSO₄ into the HDPE, the CoF of pure HDPE was substantially reduced from 0.25 to 0.10 across all concentrations. Differences in friction behaviour for the three composites were also observed in the initial metres, but later stabilised. The HDPE + 10 wt.% BaSO₄ sample demonstrated the lowest friction, with the CoF ranging between 0.06 and 0.10. In contrast, the other two radiopaque samples, HDPE + (20 and 30) wt.%, started at a slightly elevated CoF of 0.12 and 0.11, respectively, before decreasing to final coefficients of ca. 0.10. Nevertheless, all the radiopaque composites exhibited a CoF similar to that of the pure UHMWPE, indicating that the wear markers are likely to feature a similar tribological behaviour compared to the bulk inlay material.

**Figure 12.**
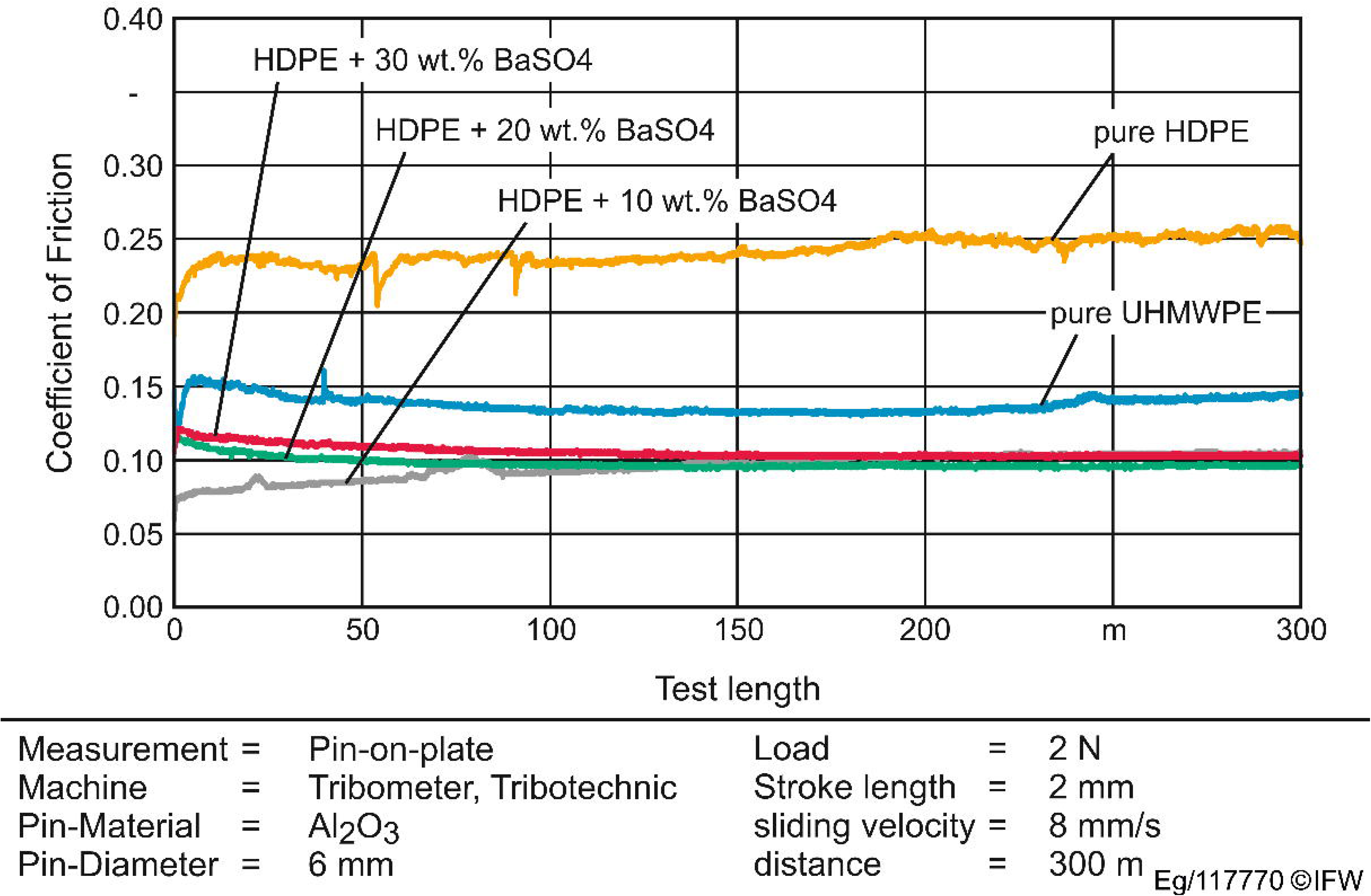
Frictional properties of the pure and radiopaque samples measured during tribometer test.

Representative surface top-view images and cross-sectional profiles of the samples after tribo-testing were analysed (Figure 13). The profiles indicate the material loss from the wear track of the samples following tribological tests. Pure HDPE demonstrated substantially higher abrasive wear than pure UHMWPE. The latter merely exhibited minor detectable scratches. Upon incorporating BaSO_4_, a shift in the wear mechanism rather towards plastic deformation than abrasion was evident in the surface images. For a BaSO_4_ concentration of 20 wt.% and higher, the roughness of the sample decreased, as indicated by the depth profile. Furthermore, the wear resistance of HDPE improved, reaching levels comparable to those of pure UHMWPE.

**Figure 13.**
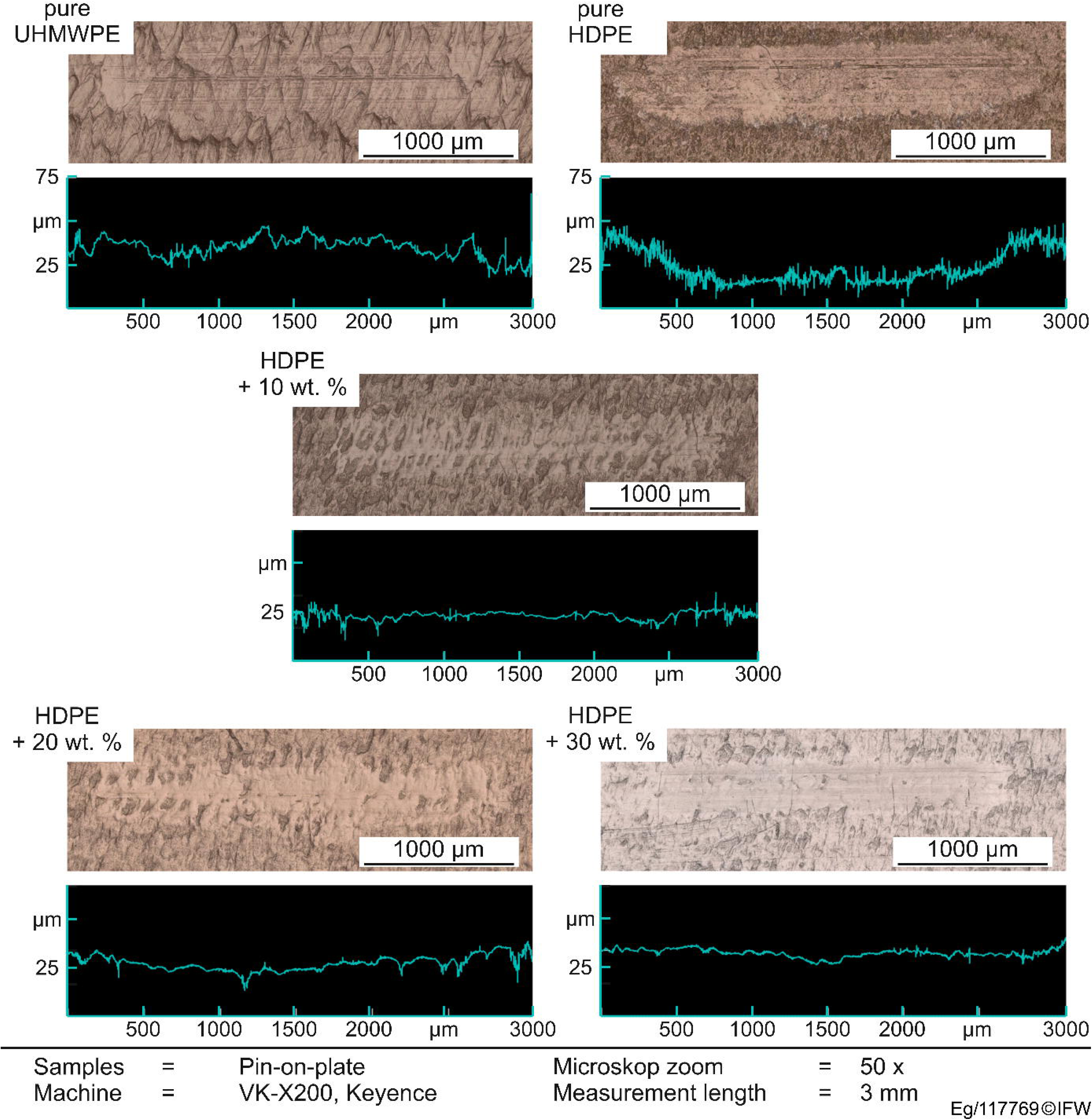
Surface image and profile section along the wear track for the samples after tribometer test.

## 4. Discussion

The present study aims to address inlay wear, one of the common causes of TKA failure. Presently, there are no standardised clinical methods for evaluating inlay wear *in vivo* outside routine radiological monitoring, which can be inaccurate in predicting the extent of wear damage [33]. This study confirms the basic feasibility of integrating wear markers into the articulating inlay surface. This was achieved by micro-milling pre-defined geometric patterns, comprising holes and/or grooves, on the surface of milled PE1000 inlays. These microstructures were strategically positioned in inlay zones most susceptible to wear. The microstructures were then filled with a biocompatible radiopaque polymeric composite using pellet extrusion.

As part of the initial parameter studies for micro-milling the cavities, 450 parameter combinations were examined on a flat UHMWPE sample using burr formation as the quality criterion to eliminate additional manufacturing steps. We were not able to produce completely burr-free samples for any of the parameter combinations. In contrast, at full tool width, burr-free samples were produced in a previous study, with the following process parameters: tool diameter D_T_ = 0.8 mm, cutting edges z = 2, cutting speed v_c_ = 25.13 m/min, feed per tooth f_z_ = 0.075 mm, and cutting depth a_p_ = 0.1 mm using conventional slot milling [24]. In the present study, the same set of parameters resulted in an average burr class of 4.00, characterised by multiple threads. The observed discrepancy could be attributed to the difference in the number of cutting edges of the tools used. In this study, tools with z = 3 were used, while the previous study used z = 2. This suggests that a larger number of cutting edges increases burr formation, although this has not yet been proven or reported in literature. Furthermore, no statistically significant regression of the experimental results could be determined, which suggests that no further parameters for optimising the process could be determined. At the same time, no high standard deviations in the average burr results were observed, indicating a stable process. This could be due to the technique used to evaluate the burrs.

The manual evaluation of burr formation using microscopic images involves a subjective component that can lead to inaccuracies. Comparable works, such as those by Bajpai et al. [34], quantified burr formation based on burr height and width in Ti6Al4V samples. Such a quantification was not possible in the present work because of the material properties of PE1000, specifically its high reflectivity, making the use of optical measurement methods challenging. A future study with quantitative burr determination could enable regression and thus provide optimal process parameters.

In the context of micro-structuring on the standard inlays, increased burr formation was observed compared to the parameter studies for the same parameters. The defined parameters resulted in an average burr class of 2.67 on flat surfaces, while micro-cavity production on free-form surfaces showed an average burr class of 4.66 (Table 6). The increased burr formation indicates inconsistent process parameters (cutting depth and width), which cannot be realised owing to the marker cavity alignment on free-form surfaces [35]. The alignment of tool and material, as exemplified by the cavities that align with the inlay backside, intensified this circumstance. However, the results from Table 6 contradict this assumption, since similar burr formation existed for both alignments. This indicates that geometric properties are not the primary cause. An alternative explanation could involve machine kinematics. In particular, five-axis manufacturing could lead to a more unstable process due to additional degrees of freedom. A comparison of different machines, considering the axis and position accuracy, could provide further insights. A process-parallel simulation of the machine axes is a promising approach [36].

During the final contouring of the inlay surface, it was also found that some of the markers had been dislodged from the microstructures. This can be attributed to the manual filling process of the marker cavities, indicating an insufficient material bond and form fit between the marker material and the inlay material. The phenomenon occurred independently of the structural orientation and alignment, but not in the case of drill holes. An investigation of the flow behaviour of the marker material could provide further insights and help to adjust the extrusion parameters. In addition, X-ray examinations showed that the cavities were often not completely filled, which can lead to a reduced static friction. Therefore, further investigations on the introduction of markers into the cavities are necessary. In addition, a mechanical and automated filling process may provide more reproducible results.

In a preliminary study, it was shown that a minimum marker concentration of 20 wt.% BaSO₄ was sufficient to achieve radiographic visibility of the markers within the inlay [24]. Despite all the radiopaque composites used in the present study exhibiting a substantially higher radiopacity than the pure samples, the attenuating effects of the soft tissue will reduce their intensity *in vivo* [37]. To compensate for this attenuation, a minimum concentration of 20 wt.% BaSO₄ was thus considered sufficient.

The X-ray investigations showed differences in the visibility of the projected marker shapes. Although the holes offered manufacturing advantages, their visibility was lower than that of the grooves. In particular, grooves that were orthogonal to the X-ray direction showed lower visibility because less material attenuated the radiation. Grooves aligned in the direction of radiation provided the best visibility. However, adjustments to the marker material could lead to manufacturing aspects being given more weight in the choice of the marker geometry. An anterior offset of the patterns, as was the case in P9, could enable spatially resolved wear determination and allow a more detailed evaluation of the wear behaviour. This could theoretically make it possible to carry out an isolated revision of the inlay, with personalised inlays adapted to the patient’s inlay wear patterns. This would be based on a more extensive study of the wear behaviour of inlays with integrated markers. In addition, the examination of patterns with combined orientations (AP and lateral) could provide new insights into the use of multiple X-ray views.

For this study, it was essential that the wear markers exhibited wear characteristics similar to those of the bulk inlay to accurately reflect the actual wear occurring on the inlay surface. Incorporating the different contrast agent concentrations to the pure HDPE reduced its CoF by 60% and improved its wear resistance to levels comparable to that of pure UHMWPE. The tribological investigations showed that adding different BaSO_4_ concentrations produced similar friction properties. It is also important to note that the UHMWPE sample used in this study had a higher roughness than that of commercial inlays [38] and was neither crosslinked nor sterilised, which might lead to wear characteristics differing from conventional inlays.

A limitation of this study is that wear markers were observed only in AP radiographic views. For clinical application, both AP and lateral views would be necessary to accurately visualise the extent of wear. Additionally, owing to the challenging processability of UHMWPE, HDPE was utilised, which is not typically employed in modern TKA. Future studies should optimise material selection to ensure clinical relevance. An in vitro investigation of the wear behaviour of the marker-integrated inlays (i.e. ISO 14243) will provide further insights into the suitability of the marker material for use in a TKA. However, the results obtained thus far are promising with regard to the tribological properties.

## 5 Conclusions

We demonstrated the basic feasibility of integrating wear markers into the TKA inlay. The micro-milling approach allowed the production of varying marker geometries at different positions and orientations on the inlay surface. The vertical grooves and drilled holes showed the highest potential for cavities for wear markers. Nevertheless, more work is required to define optimal process parameters for the production of burr-free cavities. A future parameter study with quantifiable measurement variables is proposed. A major challenge to the production of the marker cavities was the persistent burr formation, which suggested that the manufacturing five-axis process was unstable. A study with process-parallel simulation of the axis kinematics is proposed to determine the influence of local engagement conditions on burr formation. The random dislodgement of markers during final contouring revealed an insufficient material bond between the marker and inlay. The inadequate filling of the cavities could also be demonstrated based on radiographic evaluation. Accordingly, there is still a gap in research in marker extrusion, which should be investigated in future studies on automated filling. A further investigation of the wear behaviour and measurability of the markers in the context of in vitro experiments will provide insights into marker ablation and indirect wear measurement based on the change in the wear marker depth. An anterior offset of the patterns could provide information on localised wear at specific inlay locations. This could enable personalised implant regeneration using custom implant components. Furthermore, it would be interesting to analyse the cytotoxicity of the wear particles to rule out contrast-induced biological reactions. Nonetheless, this study introduces a novel technique with the potential of facilitating the timely detection of TKA inlay wear *in vivo*, a challenge to the longevity of joint replacements.

## Data Availability

All data produced in the present study are available upon reasonable request to the authors

https://service.tib.eu/trr298-repository/dataset/radiopaque-inlay-markers

## 6 Acknowledgements

The authors wish to express their gratitude to Madina Shamsuyeva and Dijan Iliew from the Institute for Plastics and Circular Economy, Leibniz Universität Hannover, who fabricated and supplied the radiopaque composites; Gerrit Hohenhoff and Matthias Henzler from the Laser Zentrum Hannover^2^, Germany for providing their technical knowledge and conducting the material extrusion and to Abinadab Sinnathurai for his contribution to the experiments and evaluation of the experimental data.

